# Heterogeneous ageing of brain networks across the Alzheimer’s disease continuum

**DOI:** 10.1101/2025.05.14.25327582

**Authors:** Yanxi Huo, Weijie Huang, Zhengzhao Liu, Tianyu Bai, Haojie Chen, Yichen Wang, Kexin Wang, Daoqiang Zhang, Jian Cheng, Yu Sun, Guolin Ma, Cui Zhao, Zhanjun Zhang, Ni Shu

## Abstract

Accelerated brain ageing has been implicated in Alzheimer’s disease (AD), which is shaped by genetic risk, molecular pathology, and disease processes. However, the spatial heterogeneity of brain ageing patterns across different functional networks along the AD continuum remained largely unexplored. In the present study, we developed a network-specific brain age prediction model trained on structural MRI from 28,341 healthy participants. By applying model to cross-sectional and longitudinal data from ADNI cohort, we estimated predicted age differences (PADs) and their change rates at individual level. Distinct network-specific brain ageing trajectories with disease progression were identified. Progressive MCI individuals showed early PAD deviations in the default mode network and accelerated changes in attention networks. Network-wise PAD dynamics mediated the effect of AD genetic risk and pathology on cognitive decline. Finally, integrating PAD features can improve predictive accuracy of MCI-to-AD conversion (AUC = 0.95). These findings highlight network-specific brain age PAD as sensitive biomarkers for early detection and monitoring of individualized AD risk.

## Introduction

The human brain undergoes progressive structural and functional degeneration with increasing age^1^. Accurately mapping the age-related trajectories of these changes is essential for understanding the normal and pathological ageing processes. Recently, brain age prediction models based on structural magnetic resonance imaging (MRI) of large samples of healthy individuals have gained widespread attention, as a promising tool for quantifying brain health at the individual level^2–4^. From these models, brain age can be non-invasively estimated, with the difference from chronological age—termed the predicted age difference (PAD)—serving as a dimensional index of deviations from normal ageing^5^.

PAD has shown great promise in identifying individuals at high risk of neurodegenerative diseases, particularly within the Alzheimer’s disease (AD) spectrum. Elevated PAD, indicative of advanced brain ageing, has been associated with multiple AD-related risk factors, including amyloid pathology, apolipoprotein E (APOE) ε4 carrier status, and cognitive impairment^6–9^. Our prior research also demonstrated accelerated brain aging in amnestic MCI, which was related to individual cognitive decline, risk factors for AD and clinical progression^10^, reinforcing the utility of PAD as a sensitive, non-invasive imaging biomarker for early diagnosis and clinical monitoring of brain disorders.

Beyond global brain ageing patterns, spatial heterogeneity of neurodegeneration is important for understanding the pathophysiology of AD^11,12^. AD pathology unfolds in a characteristic cascade— beginning with amyloid-β (Aβ) deposition, followed by tau-associated neuronal injury, and ultimately resulting in widespread neuronal loss and brain atrophy^13^. With neuroimaging techniques, previous studies have depicted the neurodegenerative pattern across the AD continuum. These changes follow a specific spatiotemporal sequence, initially affecting medial temporal structures such as the entorhinal cortex and hippocampus^14^, and gradually extending to large-scale functional brain networks, including the default mode network (DMN), limbic system, and frontoparietal control network^15^. Compared with unimodal brain networks, these transmodal networks are more vulnerable to pathological ageing^16–18^. However, most recent brain age models only compress high-dimensional brain imaging data into a single global metric, overlooking heterogeneous spatial patterns of brain ageing^19^.

Additionally, most brain age studies focus on the cross-sectional data, while longitudinal analysis can provide key advantages for capturing the dynamic nature of brain ageing^20,21^. This is particularly relevant for the prodromal stages of AD, where clinical manifestations and disease progression vary substantially among individuals, even within the same diagnostic stage^22^. Prior studies have reported that patients with progressive mild cognitive impairment (pMCI) tend to show greater PAD at baseline than those with stable MCI (sMCI)^10,23^. However, cross-sectional analysis can only provide static snapshots and cannot determine whether the observed differences are due to an initially elevated PAD or to accelerated ageing over time. The longitudinal trajectory of PAD changes among individuals at high risk for AD has remained largely unexplored.

Thus, in the present study, we proposed a network-specific brain-age prediction framework and calculated PADs across seven functional networks at the individual level. The prediction models were applied to a longitudinal cohort across the AD continuum. A nested case–control design was used to characterize the longitudinal temporospatial trajectories of brain ageing across diagnostic subgroups. Furthermore, we examined the relationships between network-specific brain ageing dynamics and genetic risk, molecular pathology of AD and cognitive performance, to explore potential links between accelerated brain ageing and disease mechanisms. Finally, we evaluated the effectiveness of network-wise brain age markers in supporting early prediction of clinical conversion of patients with MCI.

## Results

### Experimental outline and baseline characteristics

We implemented a multistage analytical framework to investigate network-level brain ageing patterns and their clinical relevance across the AD continuum (Fig. 1). Using a large-scale healthy ageing reference sample from the UK Biobank (*n* = 28,341; age range 44–83 years), we trained and validated network-specific brain age prediction models (Fig. 1a, b). These models were then applied to an independent cohort from the Alzheimer’s Disease Neuroimaging Initiative (ADNI; *n* = 1,478), comprising 542 cognitively normal participants, 655 individuals with MCI, and 281 patients with AD (Fig. 1c). This enabled external validation and individual-level PAD assessment across networks. The demographic details of both datasets are provided in Supplementary Table S1.

**Fig. 1.**
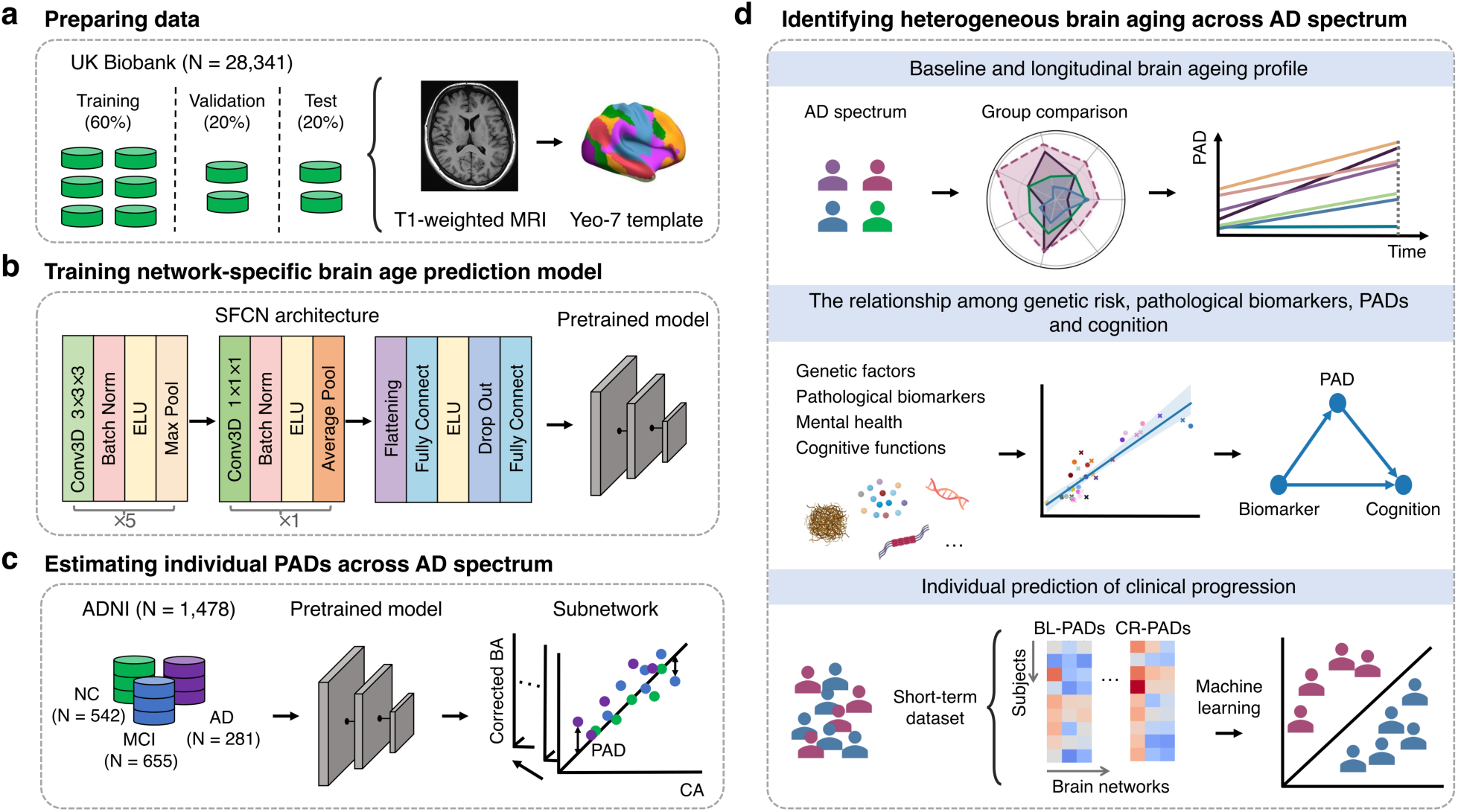
Schematic outline of the network-specific brain age analysis pipeline. **(a)** Participants from the UK Biobank (N=28,341) were randomly divided into training (60%), validation (20%), and test (20%) sets. Network-based features were extracted from T1-weighted structural images on the basis of the Yeo-7 network template. **(b)** For each network, an SFCN model was trained on the UK Biobank training and validation sets to predict brain age. **(c)** The trained model was applied to the ADNI cohort (N=1,478) to estimate network-specific brain ages and calculate corresponding PADs for each participant. PADs were then linearly corrected to eliminate the effect of chronological age (gray dots: NC; blue dots: MCI; purple dots: AD). **(d)** Longitudinal analyses were conducted to explore heterogeneity in brain ageing patterns across the AD spectrum and the temporal evolution of subnetworks. Statistical analyses were performed to elucidate the relationships among genetic risk, pathological biomarkers, PADs, and cognition. Machine learning was utilized to evaluate the predictive power of baseline PAD and its annual rate of change, derived from a short-term dataset, in forecasting long-term conversion to AD. AD, Alzheimer’s disease; ADNI, Alzheimer’s disease Neuroimaging Initiative; Average Pool, average pooling layer; BA, brain age; Batch Norm, batch normalization layer; BL-PADs, baseline PADs; CA, chronological age; Conv3D, 3D convolutional layer; CR-PADs, change rate of PADs; ELU, exponential linear unit; Fully Connect, fully connected layer; Max Pool, max pooling layer; MCI, mild cognitive impairment; NC, normal control; PAD, predicted age difference; SFCN, simple fully convolutional network.

To depict the spatiotemporal pattern of brain ageing across the AD spectrum, a nested case‒control approach was conducted using longitudinal data from the ADNI cohort (Fig. 1d). On the basis of diagnosis at baseline and clinical progression during follow-up, participants were categorized into the following four subgroups: normal controls (NCs, *n* = 189), sMCI patients (*n* = 300), pMCI patients (*n* = 157), and AD patients (*n* = 276) (see Method 2). The detailed subgrouping criteria are described in the Methods section (see Method 5). The patient demographics and clinical profiles are provided in Table 1.

**Table 1.** Baseline characteristics of ADNI participants stratified by clinical diagnosis over time.

| Characteristics | NC, *n*=189 | sMCI, *n*=300 | pMCI, *n*=157 | AD, *n*=276 | *H*/$\chi^2$ | *P*-value |
| --- | --- | --- | --- | --- | --- | --- |
| Visits | 743 | 916 | 425 | - | - | - |
| Visits per participant | 4.00 (3.00, 5.00) | 3.00 (2.00, 4.00) | 2.00 (2.00, 3.00) | - | - | - |
| Interval, years | 1.50 (1.03, 1.83) | 1.05 (1.00, 1.50) | 1.02 (1.00, 1.30) | - | - | - |
| Age at baseline, years | 74.40 (71.20, 76.40) | 73.15 (68.50, 76.55) | 72.90 (68.60, 76.40) | 73.90 (70.10, 78.05) | 9.159 | 0.027 |
| Sex, female (%) | 87 (46.00%) | 178 (59.30%) | 91 (58.00%) | 151 (54.70%) | 8.983 | 0.030 |
| Education, years | 16.00 (14.00, 18.00) | 16.00 (14.00, 18.00) | 16.00 (14.00, 18.00) | 16.00 (13.00, 18.00) | 14.708 | 0.002 |
| APOE ε4 carriers | 55 (29.10%) | 128 (42.70%) | 103 (65.60%) | 199 (72.10%) | 113.600 | $1.84 \times 10^{-24}$ |
| PHS | -0.17 (-0.43, 0.48) | 0.26 (-0.21, 0.95) | 0.72 (0.10, 1.49) | 0.91 (0.38, 1.53) | 128.283 | $1.27 \times 10^{-27}$ |
| CSF Aβ<sub>42</sub>, pgml<sup>-1</sup> | 1420.63 (917.91, 1742.77) | 723.50 (539.80, 1001.95) | 638.30 (541.80, 724.28) | 545.30 (428.55, 708.09) | 72.228 | $1.42 \times 10^{-15}$ |
| CSF tTau, pgml<sup>-1</sup> | 236.39 (191.10, 268.89) | 240.60 (181.25, 370.70) | 347.05 (264.10, 453.25) | 342.76 (269.03, 460.86) | 44.908 | $9.68 \times 10^{-10}$ |
| CSF pTau181, pgml<sup>-1</sup> | 20.87 (16.76, 25.31) | 21.95 (15.96, 37.53) | 34.72 (26.35, 48.15) | 33.71 (25.88, 45.24) | 50.645 | $5.82 \times 10^{-11}$ |
| Glucose metabolism | 7.81 (6.11, 10.11) | 11.51 (8.46, 16.18) | 15.43 (12.30, 19.04) | 22.38 (17.51, 30.24) | 99.132 | $2.39 \times 10^{-21}$ |
| Tau-IT | 1.30 (1.23, 1.33) | 1.31 (1.27, 1.40) | 1.89 (1.44, 2.03) | 1.76 (1.51, 1.89) | 34.963 | $1.24 \times 10^{-7}$ |
| Tau-ENT | 1.15 (1.09, 1.22) | 1.20 (1.11, 1.35) | 1.51 (1.49, 1.66) | 1.44 (1.31, 1.53) | 26.686 | $6.85 \times 10^{-6}$ |
| Tau-metaROI | 1.24 (1.18, 1.26) | 1.25 (1.19, 1.31) | 1.86 (1.40, 1.90) | 1.72 (1.36, 1.80) | 32.552 | $4.00 \times 10^{-7}$ |
| GDS | 0.00 (0.00, 1.00) | 1.00 (1.00, 2.00) | 2.00 (1.00, 3.00) | 1.00 (1.00, 3.00) | 69.785 | $4.75 \times 10^{-15}$ |
| NPI | 0.00 (0.00, 0.00) | 1.00 (0.00, 3.00) | 2.00 (0.00, 4.00) | 3.00 (2.00, 7.00) | 123.270 | $1.52 \times 10^{-26}$ |
| MMSE score | 30.00 (29.00, 30.00) | 29.00 (27.00, 30.00) | 26.00 (25.00, 28.00) | 22.00 (19.00, 25.00) | 489.572 | $8.68 \times 10^{-106}$ |
| MoCA score | 27.00 (26.00, 29.00) | 23.50 (22.00, 25.00) | 19.00 (17.50, 20.25) | 15.00 (11.25, 19.75) | 69.449 | $5.60 \times 10^{-15}$ |

Continuous data are described as median (interquartile range) and categorical variables are presented as numbers (percentages). Differences across different groups were compared using Kruskal–Wallis rank-sum tests for continuous variables and Pearson’s Chi-square tests for discrete variables. “Visits” denotes the total number of clinical assessments per group. “Visits per participant” refers to the average number of assessments per individual. “Interval” indicates the mean duration (in years) between consecutive assessments. AD, Alzheimer’s disease; Aβ , β-amyloid; CSF, cerebrospinal fluid; ENT, entorhinal; GDS, Geriatric Depression Scale; Glu, glucose; IT, inferior temporal; MCI, mild cognitive impairment; sMCI, stable MCI; pMCI, progressive MCI; MMSE, Minimum Mental State Examination; MoCA, Montreal Cognitive Assessment; NC, normal control; NPI, Neuropsychiatric Inventory; PHS, polygenic hazard score; pTau181, phosphorylated tau 181.

### Network-specific brain age prediction modelling

With respect to the UK Biobank training set, we developed network-specific brain age prediction models. For each of the Yeo-7 networks^24^, an enhanced simple fully convolutional network (SFCN) model was used to predict network-specific brain age from MRI-derived imaging features. The SFCN framework integrates efficient feature extraction with a minimally parameterized fully convolutional neural network architecture^25^, allowing for precise characterization of age-related brain structural changes and spatially distinct ageing patterns. Details of the network architecture are provided in Method 1. Before calculating the PAD, linear correction was performed to eliminate the effect of chronological age on brain age (see Method 4).

These models demonstrated robust performance in brain age prediction. In the UK Biobank test set, network-specific predictions yielded Pearson correlation coefficients (r) ≥ 0.87, coefficients of determination (R^2^) ≥ 0.67, and mean absolute errors (MAEs) < 3.60 (Fig. 2a). In the ADNI cohort, the performance of the predictions in the cognitively normal group further validated the strong generalization ability across independent datasets, with r ≥ 0.84, R^2^ ≥ 0.55 and MAE < 2.66 (Fig. 2b). The comprehensive results are presented in Supplementary Table S2.

**Fig. 2.**
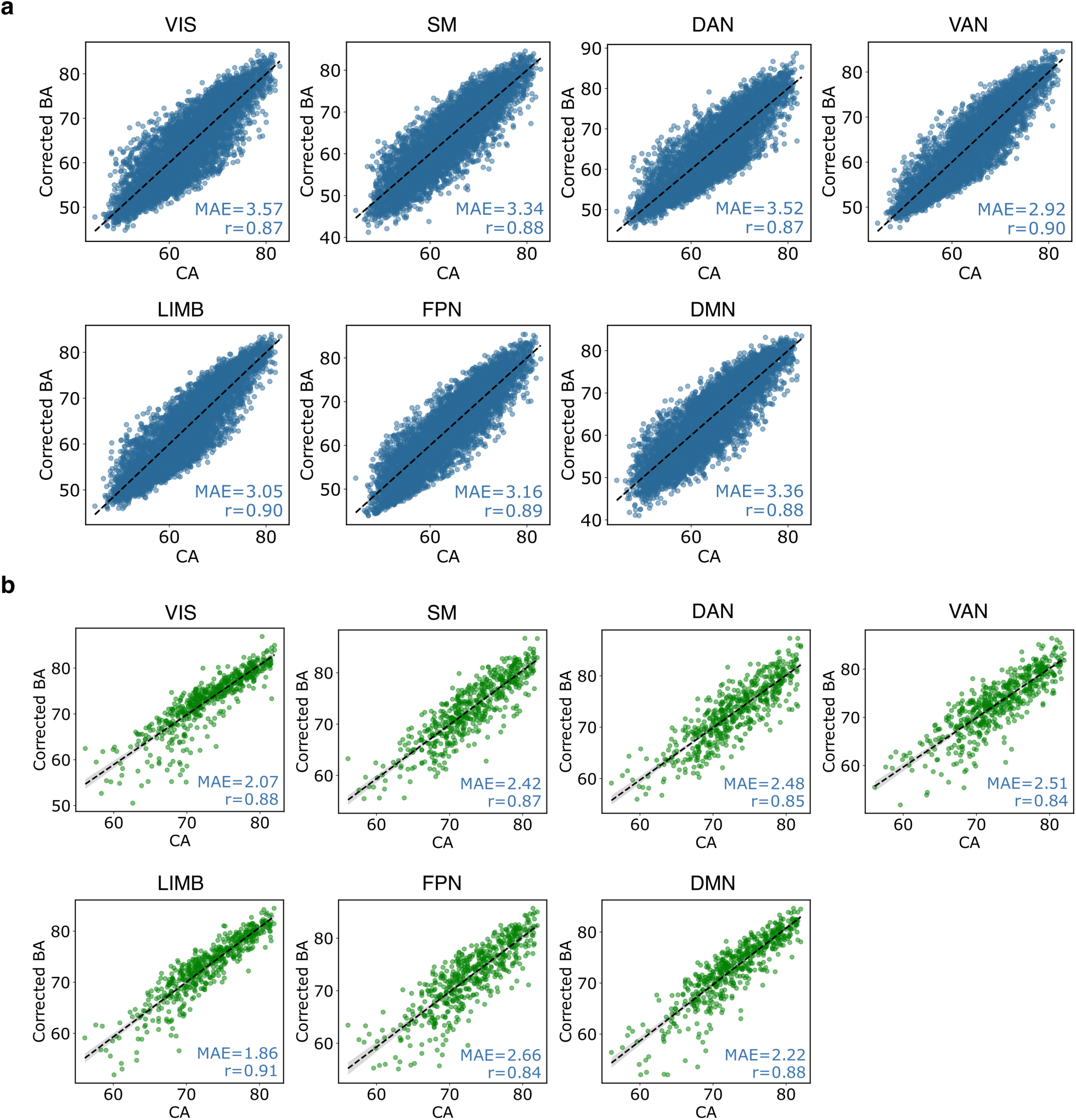
Performance of network-specific brain age prediction models in predicting chronological age. **(a)** Performance of the trained models on the UK Biobank test set. **(b)** Performance of the trained models on the NCs from the ADNI cohort. BA, brain age; CA, chronological age; DAN, dorsal attention network; DMN, default mode network; FPN, frontoparietal network; LIMN, limbic network; MAE, mean absolute error; NC, normal control; SMN, somatomotor network; VAN, ventral attention network; VN, visual network.

### Identifying heterogeneous brain ageing patterns across the AD spectrum

Network-level brain ageing was quantified using corrected PAD values derived for each participant in the ADNI cohort. The PAD reflects the deviation of predicted brain age (BA) from chronological age (CA), thereby indexing the degree of brain ageing at the individual level.

As shown in the top panel of Figure 3a, linear models adjusted for age, sex, education, and APOE ε4 status revealed significant differences in baseline PADs in all networks among groups (all *P* values < 0.001; Supplementary Table S3). The estimated marginal means indicated that the AD group consistently presented the highest PADs (range: 2.16–2.97), followed by the pMCI group (range: 1.57– 2.27), whereas the sMCI and NC groups presented considerably lower values across networks. Post hoc comparisons of adjusted marginal means, corrected for multiple comparisons using Tukey’s honest significant difference (HSD), revealed that the AD group had significantly greater PADs compared with those of all other groups across all networks, except in the visual (VN) and limbic (LIMN) networks, where differences in the pMCI group did not reach significance. The pMCI group, in turn, presented significantly greater PADs compared with those of the sMCI (all *P* values < 0.05) and NC (all *P* values < 0.001) groups. No significant differences in PADs were detected between sMCI patients and NCs in any network (Supplementary Table S4).

**Fig. 3.**
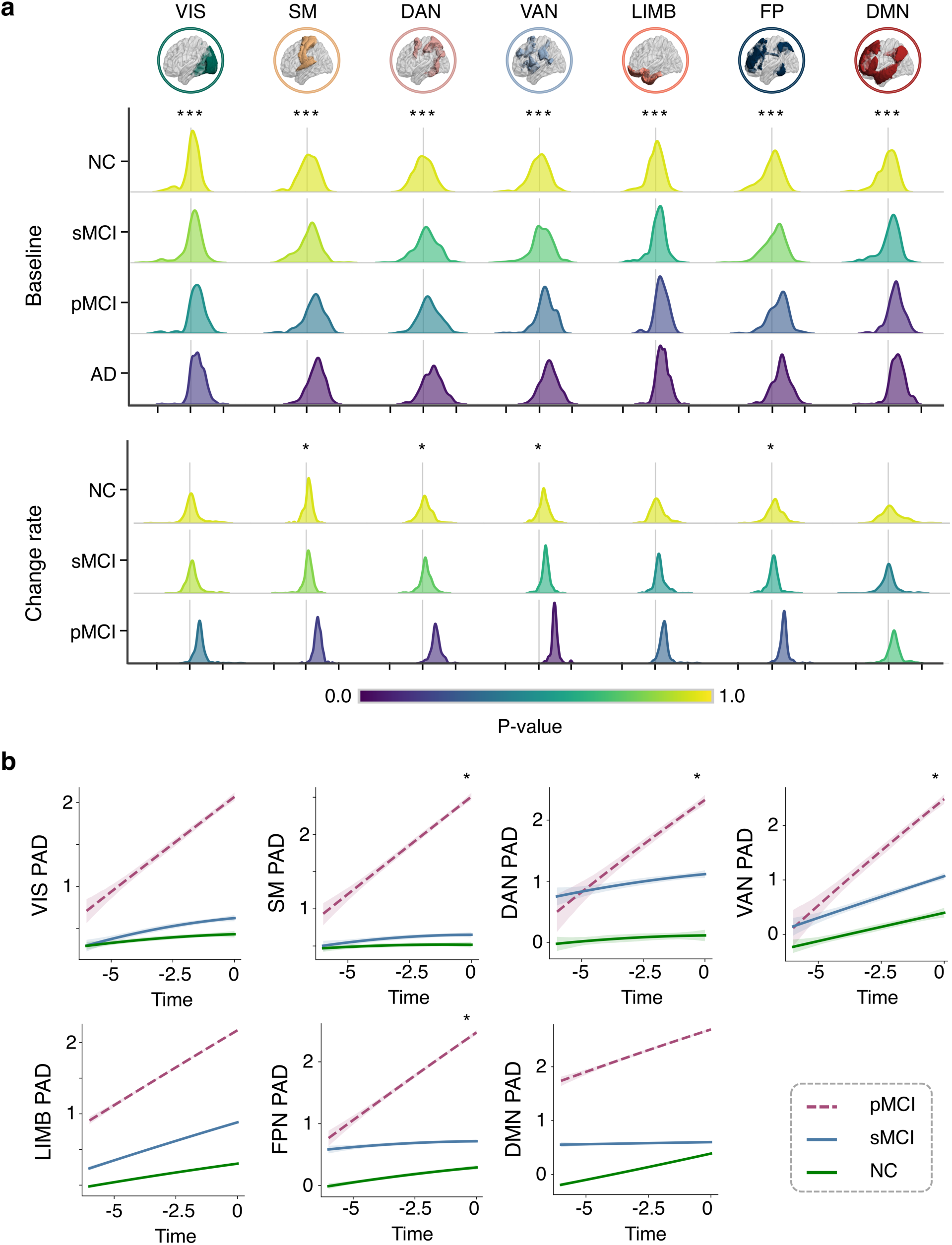
Group comparison of network-wise PAD and annual change rates of PAD across the AD spectrum. **(a)** Distribution of network-wise PADs at baseline (top) and their annual change rates before AD diagnosis (bottom) across different groups. The colours represent *P* values from Tukey’s pairwise multiple comparison test comparing each group with the NCs, following ANOVA (\*\*\**P* < 0.001, \*\**P* < 0.01, \**P* < 0.05, and no symbols *P* ≥ 0.05). **(b)** Longitudinal trajectories of network-wise PADs over time before AD diagnosis in different groups. * denotes a significant group × time interaction (*P* < 0.05, ANOVA). ANOVA, analysis of variance; DAN, dorsal attention network; DMN, default mode network; FPN, frontoparietal network; LIMN, limbic network; MCI, mild cognitive impairment; NC, normal control; PAD, predicted age difference; SMN, somatomotor network; sMCI, stable MCI; pMCI, progressive MCI; VAN, ventral attention network; VN, visual network.

With respect to longitudinal changes in network-specific PADs (Fig. 3a, bottom panel; Fig. 3b), ANOVA revealed significant group effects in the somatomotor network (SMN) (F = 6.77, *P* = 0.034), dorsal attention network (DAN) (F = 7.62, *P* = 0.022), ventral attention network (VAN) (F = 7.96, *P* = 0.019), and frontoparietal network (FPN) (F = 8.30, *P* = 0.016). Post hoc pairwise comparisons corrected by Tukey’s HSD indicated that relative to the sMCI group, the pMCI group presented significantly greater rates of PAD increase in the SMN (T = 2.41, *P* = 0.043), DAN (T = 2.44, *P* = 0.040), and FPN (T = 2.83, *P* = 0.013). Similarly, compared with the NC group, the pMCI group presented significantly greater PAD change rates in the SMN (T = 2.42, *P* = 0.042), DAN (T = 2.65, *P* = 0.023), VAN (T = 2.81, *P* = 0.015), and FPN (T = 2.41, *P* = 0.043). No significant differences in PAD change rates were found between NCs and sMCI patients in any network (Supplementary Table S5).

To characterize the temporal sequence of network-specific alterations preceding AD onset, fitted PAD trajectories from pMCI patients were aggregated into a combined model. Retrospective analysis extending up to six years before AD diagnosis (Fig. 4) revealed that the DMN exhibited the highest PAD values throughout, suggesting that it is among the earliest and most vulnerable networks in AD pathophysiology. The estimated marginal slopes of the PAD increase (Supplementary Table S6) further supported this pattern. pMCI individuals presented the most pronounced PAD growth in the VAN (*β* = 0.40), DAN (*β* = 0.36), FPN (*β* = 0.31), and SMN (*β* = 0.30). In contrast, the VN showed the slowest rate of increase, indicating relative preservation. These findings highlighted distinct trajectories of network-specific brain ageing, with accelerated changes in attentional and control networks potentially serving as early biomarkers of AD progression.

**Fig. 4.**
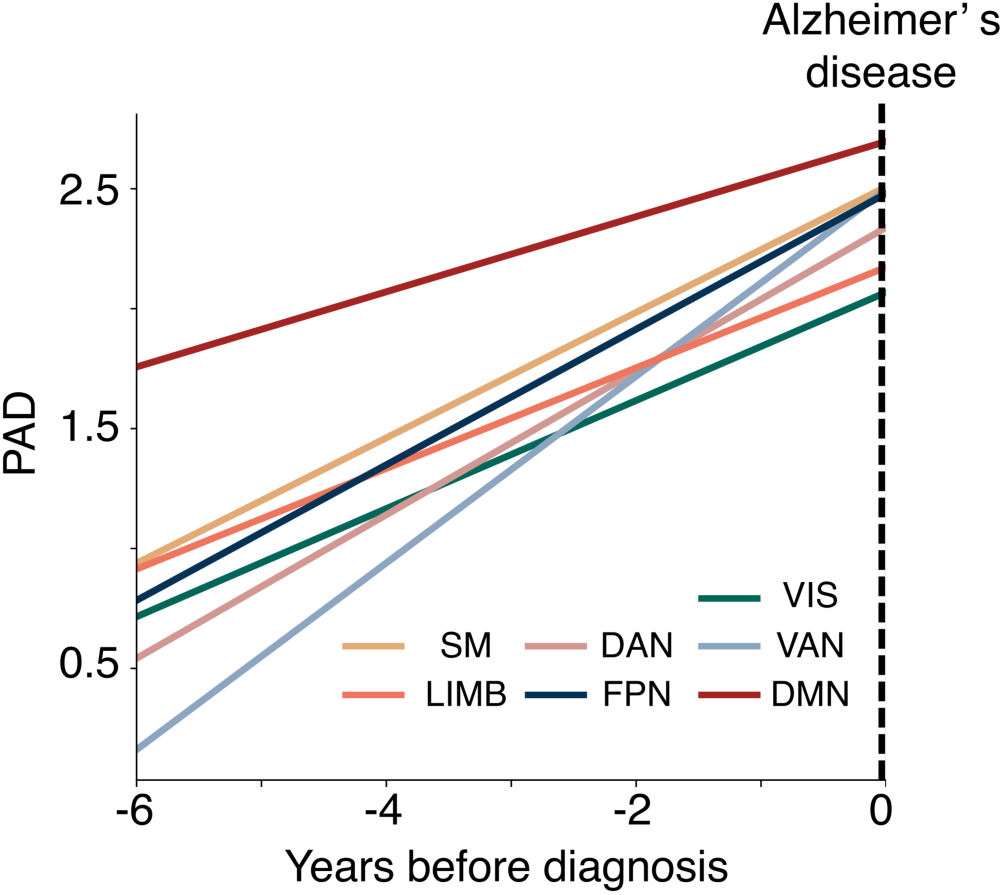
Temporal evolution of network-specific PADs before the diagnosis of AD. Year 0 represents the year of diagnosis of AD. AD, Alzheimer’s disease; DAN, dorsal attention network; DMN, default mode network; FPN, frontoparietal network; LIMN, limbic network; PAD, predicted age difference; SMN, somatomotor network; VAN, ventral attention network; VN, visual network.

### Associations with genetic, pathological, psychological, and cognitive factors

The associations of both baseline and longitudinal changes were investigated in network-wise PADs with genetic predispositions, pathological biomarkers, mental health, and cognitive functions in individuals with MCI (Fig. 5).

**Fig. 5.**
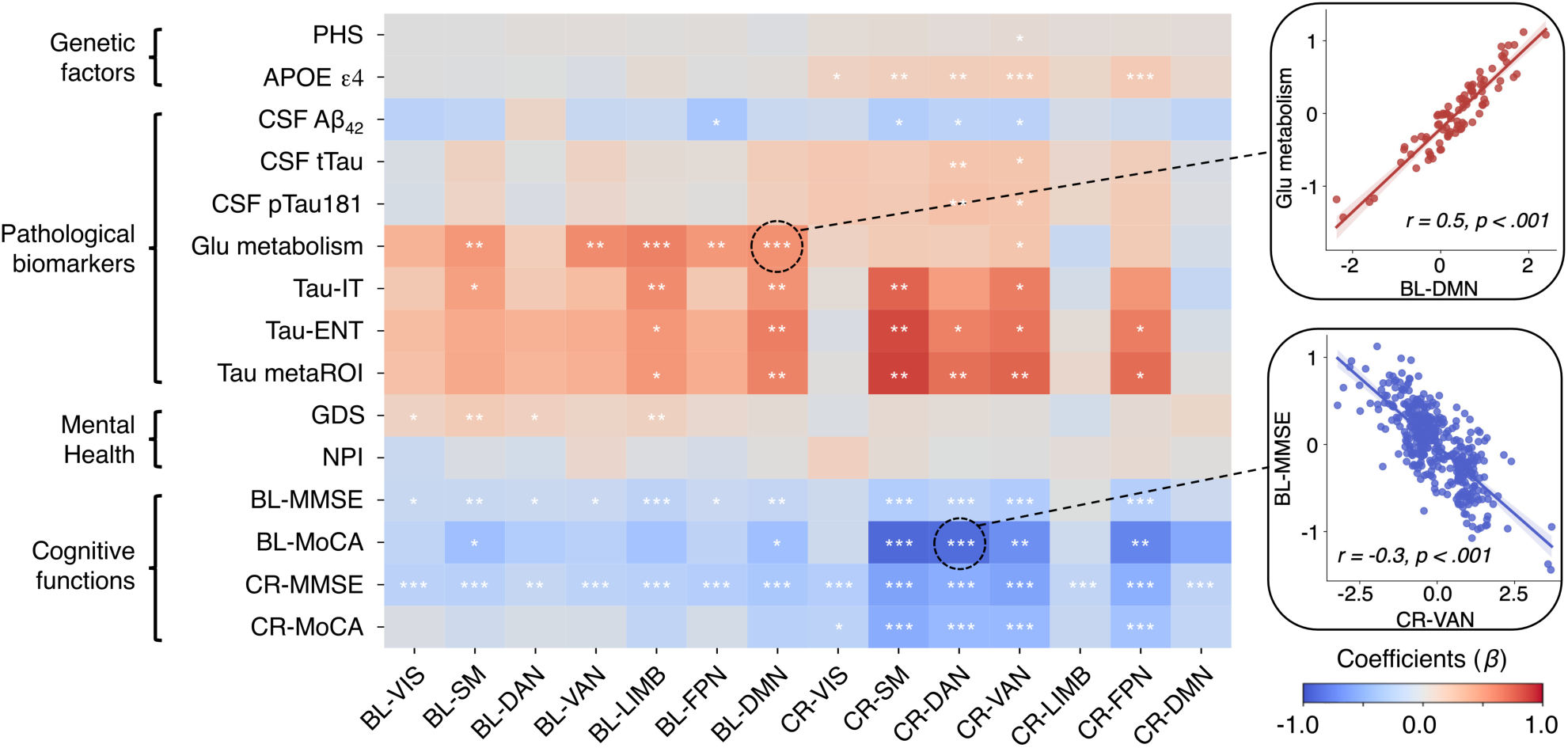
Associations of PADs with genetic risk, pathological biomarkers, and cognition. Heatmap showing the correlations of network-specific PADs at baseline (left) and their longitudinal change rates (right) in individuals with MCI with genetic factors, pathological biomarkers, mental health, and cognitive function. The colours represent beta estimates, and the asterisks denote statistically significant associations after Benjamini–Hochberg correction (\*\*\**P* < 0.001, \*\**P* < 0.01, \**P* < 0.05, and no symbols *P* ≥ 0.05). Aβ, β-amyloid; BL, baseline; CR, change rate; CSF, cerebrospinal fluid; DAN, dorsal attention network; DMN, default mode network; ENT, entorhinal; FPN, frontoparietal network; GDS, Geriatric Depression Scale; Glu, glucose metabolism; IT, inferior temporal; LIMN, limbic network; SMN, somatomotor network; MCI, mild cognitive impairment; MMSE, Mini-Mental State Exam; MoCA, Montreal Cognitive Assessment; NPI, Neuropsychiatric Inventory; PAD, predicted age difference; PHS, polygenic hazard score; pTau181, phosphorylated tau 181; tTau, total tau; VAN, ventral attention network; VN, visual network.

Among the pathological biomarkers, reduced glucose metabolism and increased PET tau deposition were significantly associated with increased PADs, predominantly in the LIMN and DMN—regions that already presented elevated PADs at baseline. However, no significant associations were observed between baseline PADs and genetic factors, such as APOE ε4 status or polygenic hazard scores (PHS), across any network (Supplementary Table S7).

With respect to psychological and cognitive measures, higher Geriatric Depression Scale (GDS) scores were positively associated with increased PADs in the VN (*β* = 0.14, *P* = 0.006), SMN (*β* = 0.20, *P* = 1.11×10^4^), DAN (*β* = 0.16, *P* = 0.003), and LIMN (*β* = 0.15, *P* = 0.002), whereas cognitive performance—assessed by the Mini-Mental State Examination (MMSE) and the Montreal Cognitive Assessment (MoCA), both at baseline and over time—was consistently and negatively correlated with PADs across all networks. This inverse relationship was particularly notable in the DMN, LIMN and SMN, where lower baseline MMSE scores and faster cognitive decline were linked to greater PADs.

With respect to longitudinal PAD changes, the APOE ε4 carrier status was significantly positively correlated with PAD change rates in the SMN (*β* = 0.17, *P* = 4.56×10^4^), DAN (*β* = 0.18, *P* = 1.72×10^4^), VAN (*β* = 0.18, *P* = 1.02×10^4^) and FPN (*β* = 0.21, *P* = 1.39×10^5^). Similarly, the PET-based tau load in the inferior temporal cortex, entorhinal cortex, and predefined meta-ROI (see Method 7) was significantly positively correlated with PAD change rates in these networks. In contrast, CSF measures (Aβ_42_, tTau and pTau181) were correlated primarily with PAD change rates in the DAN and VAN (*β* range: 0.23 to 0.27). No significant associations were found between PAD change rates and psychological measures such as the GDS or Neuropsychiatric Inventory (NPI). Furthermore, the PAD change rates in these networks were negatively correlated with cognitive performance, including baseline MMSE (*β* range: –0.26 to –0.29), baseline MoCA (*β* range: –0.69 to –0.87), annual MMSE decline (*β* range: –0.51 to –0.58), and annual MoCA decline (*β* range: –0.38 to –0.52). Collectively, these findings underscored the pivotal role of network-level accelerated brain ageing in cognitive deterioration across the AD continuum.

### Network-specific PADs mediate the links between AD risk factors and cognition

We next examined whether the prospective associations between genetic and pathological biomarkers with cognitive performance are mediated by brain-related factors. To this end, mediation analyses were conducted for each risk factor–cognition pair within a path analysis framework (see Method 9). Overall, we identified multiple significant pathways through which genetic risk and AD-related pathology may influence accelerated ageing of brain networks, which in turn leads to cognitive decline (Fig. 6). These effects were primarily observed in networks showing faster ageing trajectories during the transition from MCI to AD, including the SMN, DAN, VAN, and FPN, with network-specific modulation by both genetic and pathological factors.

**Fig. 6.**
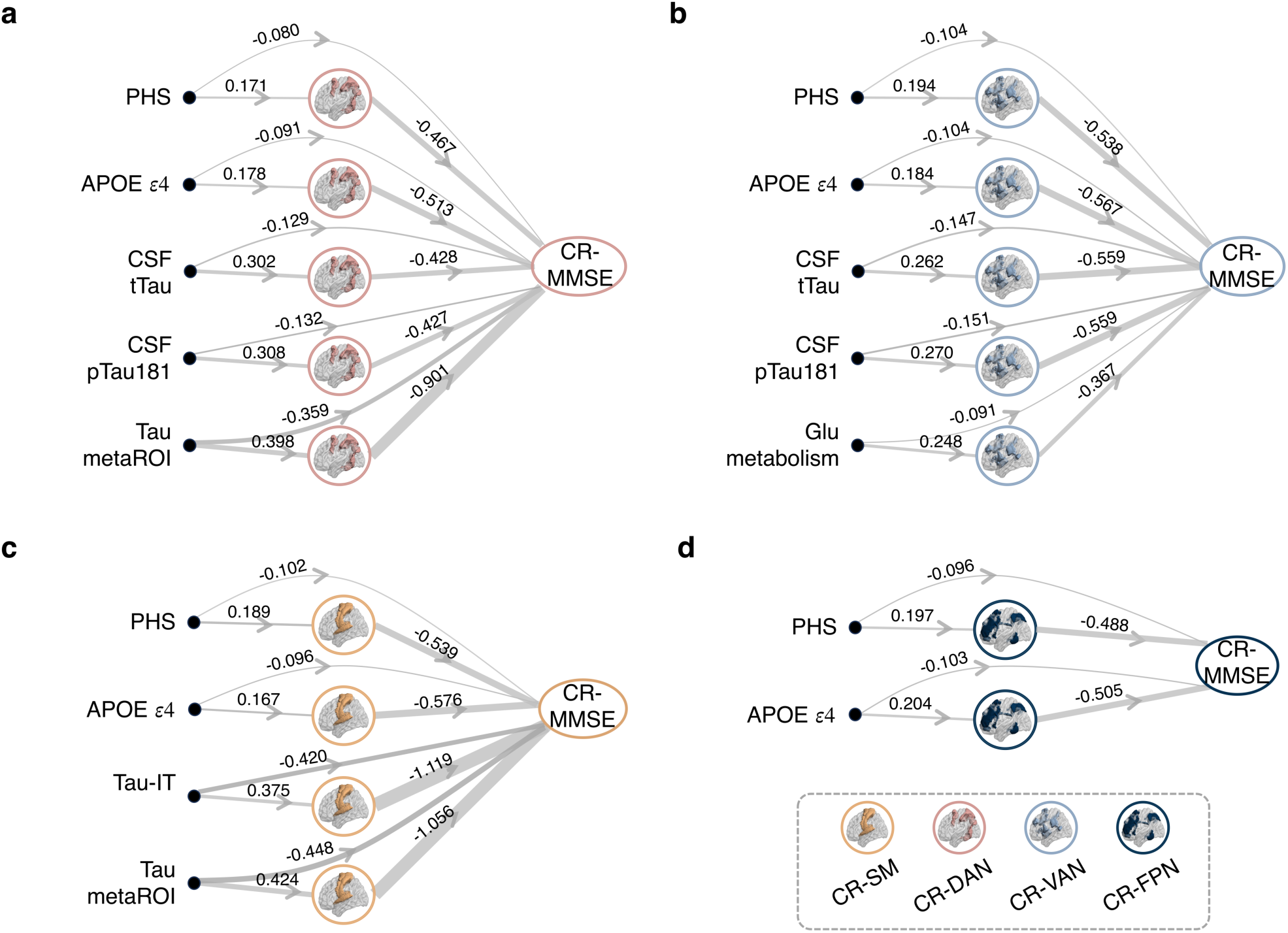
Mediating effects of PADs and the annual change rate of PADs on the relationships among genetic risk factors, pathological biomarkers and cognitive function. **a–d**, Two-step ordinary least squares regression models illustrating the pathways through which genetic factors and pathological biomarkers influence cognitive decline via annual change rates of PADs in the DAN (**a**), VAN (**b**), SMN (**c**), and FPN (**d**). Only statistically significant mediation pathways via PAD change rates are shown (*P_FDR_* < 0.05). Arrow thickness represents effect size, with numerical values indicating standardized regression coefficients. The full model statistics and confidence intervals are provided in Supplementary Table S9. CSF, cerebrospinal fluid; DAN, dorsal attention network; FPN, frontoparietal network; Glu, glucose metabolism; IT, inferior temporal; SMN, somatomotor network; MMSE, Mini-Mental State Exam; PHS, polygenic hazard score; pTau181, phosphorylated tau 181; VAN, ventral attention network.

Specifically, in individuals with MCI, PAD change rates in the SMN, DAN, VAN, and FPN significantly mediated the associations between PHS and MMSE (indirect effect range: –0.08 to –0.10; total effect = –0.24; all *P_FDR_* < 0.05), with similar effects observed for APOE ε4 (indirect effect range: –0.09 to –0.10; total effect = –0.20; all *P_FDR_* < 0.05). For pathological markers, CSF tTau and pTau181 exhibited significant indirect effects via PAD change rates in the DAN and VAN (indirect effect range: –0.13 to –0.15; total effects = –0.30 and –0.30, respectively; all *P_FDR_* < 0.05). Moreover, the PET-based tau burden (Tau-IT and tau meta-ROI) was mediated through the SMN and DAN (indirect effect range: –0.36 to –0.45; total effects = –0.63 and –0.73, respectively; all *P_FDR_* < 0.05). The statistical details are provided in Supplementary Table S9. While these results did not establish strict causality, they delineated potential directional relationships identified through established causal structure discovery methods.

### Prediction of clinical progression from MCI to AD

To evaluate the utility of network-based brain ageing features in predicting the conversion from MCI to AD, we performed feature ablation analysis using a machine learning classifier. Starting from a benchmark model based on genetic and pathological factors, we incrementally incorporated baseline PADs and their short-term longitudinal change rates (see Method 10).

As shown in Table 2 and Figure 7, among all participants diagnosed with MCI at baseline, the full-feature model—integrating genetic, pathological, and network-based ageing features—achieved the highest performance across all the metrics for identifying converters. Compared with the benchmark model, the inclusion of baseline PADs significantly improved the area under the receiver operating characteristic curve (AUC-ROC) (two-sided paired t test: T = 3.13, *P* = 0.012). Furthermore, although not significant, performance gain was observed when longitudinal PAD change rates were incorporated (T = 1.51, *P* = 0.165). Specifically, the full-feature model achieved an F1 score of 0.89 ± 0.05, a recall of 0.87 ± 0.15, an accuracy of 0.90 ± 0.05, and an area under the AUC-ROC curve of 0.95 ± 0.04, outperforming the benchmark model (F1 score: 0.74 ± 0.12; recall: 0.64 ± 0.15; accuracy: 0.79 ± 0.07; AUC-ROC curve: 0.87 ± 0.06). These results highlighted the added value of individualized network-based brain ageing markers in enhancing early AD identification and further underscored the potential of multimodal, brain-informed machine learning models for personalized risk stratification.

**Fig. 7.**
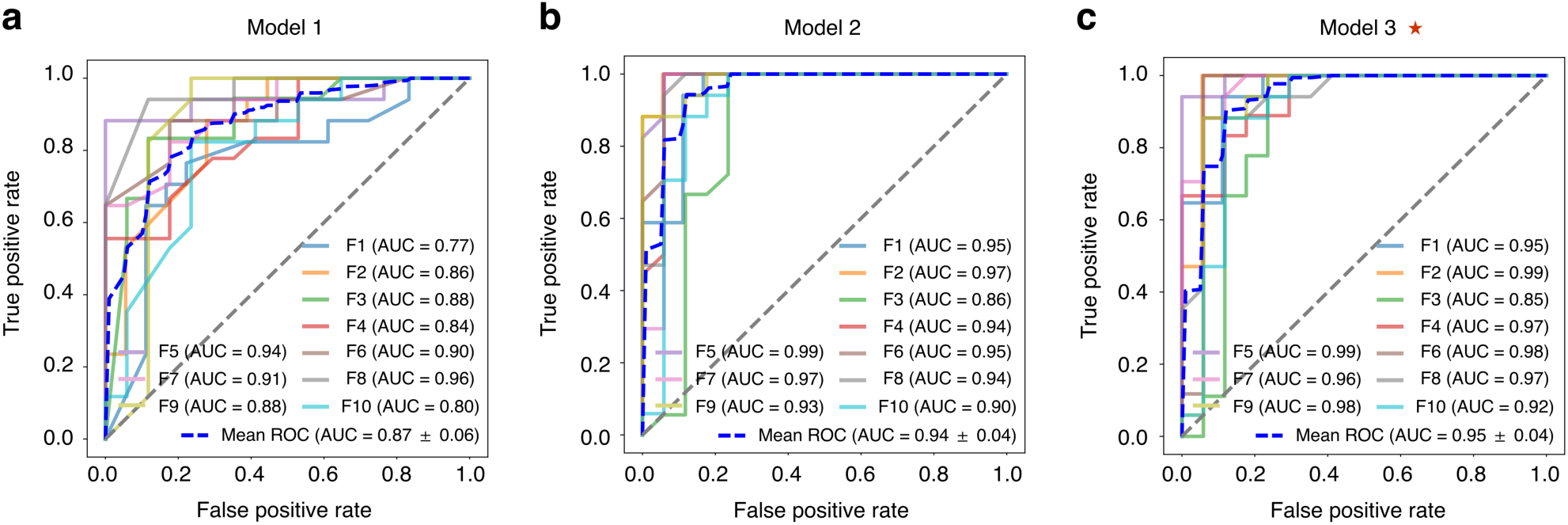
Performance of classifiers predicting mild cognitive impairment conversion to AD. **a–c**, ROC curves showing the performance of models trained for identifying individuals with MCI who later progressed to AD. Model 1 (**a**) includes only genetic and pathological biomarkers; Model 2 (**b**) incorporates additional baseline PADs; and Model 3 (**c**) further includes short-term longitudinal change rates of PADs. The red star denotes the optimal model. AD, Alzheimer’s disease; MCI, mild cognitive impairment; PAD, predicted age difference; ROC, receiver operating characteristic.

**Table 2.**
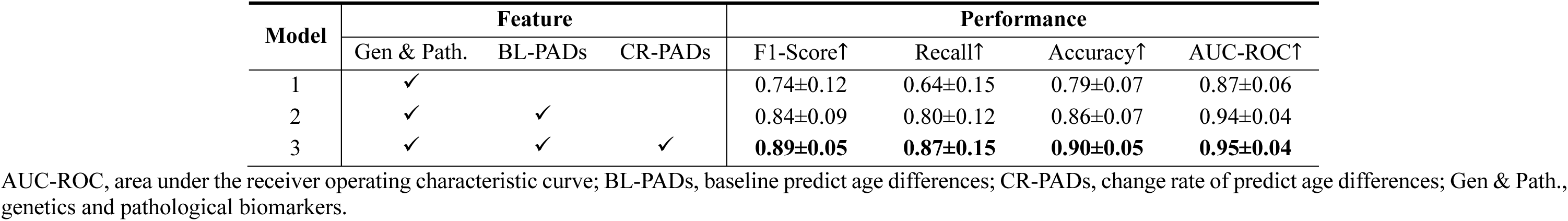
The performance of classification models.

## Discussion

In this study, we investigated the temporal evolution of network-specific brain ageing across the AD continuum, aiming to identify early vulnerable patterns and shed light on potential mechanistic links between upstream risk factors and downstream cognitive decline. Using a large-scale healthy cohort, we developed a framework for estimating the network-specific brain ages, which showed high accuracy and generalizability in external validation. We observed heterogeneous ageing trajectories of brain networks across the AD continuum. Compared with sMCI patients, pMCI patients initially presented with a higher PAD in the DMN and a faster PAD change rate in the SMN, DAN and FPN. These network-specific alterations were significantly associated with AD biomarkers. Notably, longitudinal PAD change rates proved more sensitive than baseline values, and mediated the effect between genetic factors and molecular pathology with cognitive decline. Furthermore, integrating the PAD change rate into the classification model significantly improved the predictive performance of clinical conversion in MCI patients. Together, our findings highlight network-specific brain ageing as a mechanistic and clinically informative marker for individual AD risk and disease progression.

This framework revealed a distinct pattern of heterogeneous ageing across functional brain networks along the Alzheimer’s disease (AD) continuum. Notably, progressive MCI (pMCI) individuals showed early brain-age deviations in the default mode network (DMN), followed by accelerated ageing in attention and control networks (SMN, DAN, VAN, FPN). Importantly, the longitudinal change rate of predicted age difference (PAD) emerged as a more sensitive biomarker of AD-related neurodegeneration than baseline PAD. It exhibited stronger associations with genetic risk and tau pathology, and significantly mediated their effects on cognitive decline. Incorporating network-level PAD features, particularly their temporal dynamics, substantially improved the prediction of MCI-to-AD conversion. Together, these findings highlight network-specific brain ageing as both a mechanistic pathway and a clinically informative marker for individualized risk monitoring and disease progression in AD.

To depict the spatiotemporal patterns of brain ageing in brain networks, we developed network-specific brain age prediction models. These models were trained on a large-scale dataset from the UK Biobank and were then applied directly to the ADNI without any fine-tuning. Notably, the scanning parameters of T1W MR images from the ADNI are highly heterogeneous because of the upgrade from 1.5T MR scanners to 3.0T MR scanners. Although these models were trained on images acquired from 3.0T scanners, they also demonstrated good generalization performance on images obtained from various scanners. The preprocessing of T1W MR images was minimal and included only skull stripping, normalization of nonuniform signal intensities, Talairach space transformation, removal of nonbrain tissues, and resampling. These findings suggested that the proposed framework may demonstrate high clinical utility in evaluating brain network ageing. To identify local ageing patterns, recent studies have used machine learning methods to generate saliency maps^26,27^. However, saliency maps depict regional weights of MRI features during age prediction, which may not directly reflect regional ageing patterns. In contrast, our network-specific brain age models provide a more direct and intuitive approach. In addition, our models were developed before the hypotheses were tested, which ensured that the results regarding heterogeneous brain ageing in MCI patients were independent of the way the models were built.

The results of the baseline PAD comparison indicated that the extent of brain network age deviation from actual age progressively increases with the clinical progression of patients on the AD spectrum across all networks. However, post hoc analyses did not reveal significant differences in the baseline PADs of the brain networks between the sMCI patients and NCs. These observations are consistent with findings from studies using a whole-brain brain age model^28,29^. The present findings further suggested that the increased brain age of AD spectrum patients occurs across cerebral networks. Additionally, the baseline PADs in the DMN differed the most when comparing pMCI patients with sMCI patients or NCs. These findings provide evidence that regions in the DMN are the most vulnerable to AD pathology.

From a longitudinal perspective, we revealed the network specificity of brain ageing in pMCI patients. The PAD change rates in the SMN, DAN, VAN and FPN in pMCI patients were significantly greater than those in sMCI patients and NCs, whereas the PAD change rates in the DMN did not significantly differ. These results may be due to the brain age changes following a sigmoid shape over a prolonged period^30^, suggesting accelerated ageing in the preclinical stage and subsequent slowing during clinical progression. Moreover, AD molecular pathological biomarkers spread from the DMN to other networks, such as the SMN, DAN, VAN, and FPN, leading to accelerated brain ageing in these networks. This observed spatiotemporal pattern aligns with previous fMRI studies indicating that DMN disruption precedes clinically evident symptoms and subsequently extends to networks unrelated to episodic memory, such as the executive and attention networks^31,32^, paralleling the clinical progression from memory impairments to deficits in orientation, reasoning, language, and noncognitive domains. The observed spatiotemporal patterns of brain network ageing carry important implications for understanding the progression mechanisms of AD.

In the present study, we also found that the baseline PADs, but not the PAD change rates, in the LIMN and DMN were correlated with hypometabolism and increased hyperphosphorylated tau neurofibrillary tangles in MCI patients. These findings suggested that the brain age in the LIMN and DMN may have already approached the plateau region of the sigmoid curve by the MCI stage, providing evidence that the LIMN and DMN are among the earliest networks affected by AD pathology and subsequent ageing. The DMN and LIMN function as distinct but interacting components within a memory network, collaboratively facilitating memory-based behaviours^33–35^. This finding provides the neuroanatomical basis for the memory domain being the first impaired cognitive domain in AD. Unlike baseline PAD, the PAD change rate correlated not only with glucose metabolism and tau neurofibrillary tangles but also with cerebrospinal fluid biomarkers and genetic risk factors for AD; these associations were observed predominantly in the four brain networks showing accelerated ageing in pMCI patients. These findings are consistent with previous studies reporting that CSF biomarkers are correlated with longitudinal cortical thickness loss but not with cortical thickness itself. The distinct relationships between baseline PAD and PAD change rates with different pathological markers indicate that although upstream molecular biomarkers may not directly correlate with downstream MRI-based neurodegenerative changes, they can still predict ongoing and progressive pathological processes in the cascading pathology of AD. Therefore, these network-specific brain ageing patterns provide critical neurobiological insights, clarifying the temporal and spatial interactions among different pathological biomarkers during the progression of AD.

In the present study, we used two common tools, the MMSE and MoCA, to evaluate the cognitive impairment of MCI patients. The MoCA has been shown to be superior to the MMSE in discriminating between individuals with mild cognitive impairment and those with no cognitive impairment because of its ability to detect more subtle cognitive impairment^36^. We observed that the baseline PADs of the brain networks were associated with only the baseline MMSE score and the MMSE score change rate, whereas the PAD change rates were associated with the baseline MMSE score, baseline MoCA score, MMSE score change rate, and MoCA score change rate. These findings indicated that PAD change rates can reflect more subtle cognitive impairment, thus underscoring the importance of longitudinal PAD change rates as biomarkers for AD. Combining genetic risk factors, pathological biomarkers, PADs, PADs change rates, and cognition, the present mediation analysis revealed the pathways from genetic risk factors or CSF biomarkers to accelerated ageing of brain networks to cognitive decline.

Our previous study revealed that combining baseline PAD and other AD biomarkers can predict the progression of MCI patients with an accuracy of 0.877^10^. This finding was replicated in the present study, demonstrating that incorporating baseline brain age as an additional feature alongside genetic information and pathological biomarkers significantly improves the accuracy of predicting MCI progression. Furthermore, inclusion of the PAD change rate as an additional predictor significantly improved the prediction accuracy from 0.86 to 0.90. This finding highlights the clinical importance of longitudinal monitoring of brain ageing for predicting Alzheimer’s disease risk. Overall, the proposed network-specific brain age prediction framework not only provides a deeper understanding of individual brain network ageing trajectories but also offers valuable tools for personalized prognosis and early intervention strategies in Alzheimer’s disease.

Notably, several limitations need to be considered when the present results are interpreted. First, similar to previous studies, the network-based PAD derived from our brain-age model showed a correlation with chronological age, and there is currently no consensus regarding how to address this age-related bias. Some researchers have advocated to statistically remove the effects of age from PAD values via regression methods^37^, whereas others have argued that such corrections artificially inflate model accuracy^38,39^. Therefore, developing predictive models capable of inherently minimizing or eliminating this chronological age bias remains an important direction for future research. Second, although the present study employed a longitudinal design, we only used a six-year time window and did not include participants who progressed from cognitively normal to dementia. Consequently, we were unable to accurately capture the complete trajectory of brain network ageing influenced by Alzheimer’s disease pathology. Finally, dividing the brain into only seven networks remains relatively vague, limiting the ability to detect localized brain ageing. Previous studies have also demonstrated that molecular biomarkers of AD, such as amyloid deposition and tau neurofibrillary tangles, appear mainly in specific hub regions within networks rather than uniformly across entire networks^40^. Thus, this coarse parcellation may also hinder the identification of associations between AD molecular biomarkers and localized brain ageing.

In conclusion, the present study depicted the spatiotemporal ageing of large-scale brain networks across the AD spectrum and revealed the neuroanatomical basis underlying the heterogeneous clinical progression of MCI patients from the perspective of brain network ageing. Additionally, the relationships of the PAD change rates with genetic risk factors, CSF biomarkers, cognitive functions and the clinical progression of MCI patients highlighted the importance of monitoring the longitudinal change rate of brain ageing in clinical settings.

## Acknowledgements

This research has been conducted using the UK Biobank Resource under Application Number 49749. This work was supported by the STI2030-Major Projects (2022ZD0213300, 2022ZD0211600), National Natural Science Foundation of China (82301608, 32271145, 81871425, 210510238), Open Research Fund of the State Key Laboratory of Cognitive Neuroscience and Learning (CNLZD2101, CNLZD2303), Fundamental Research Funds for the Central Universities (2017XTCX04).

## Conflict of Interest Statement

The authors declare no competing interests.

## Methods

### Method 1: study population

The magnetic resonance imaging (MRI) (see Method 2) and clinical data (see Method 6) used in the present study were obtained from 29,819 participants across two independent datasets—the UK Biobank (https://www.ukbiobank.ac.uk)^41^ and the ADNI (https://adni.loni.usc.edu/), encompassing the ADNI1&GO, ADNI2, and ADNI3 phases. The participants ranged in age from 44 to 83 years, and longitudinal data was available for 1,042 individuals. The demographic characteristics and sample sizes are presented in Supplementary Table S1. Written informed consent was obtained from all participants in their respective studies, and the protocols of this study were approved by the Ethics Committee of the Institutional Review Board at the Beijing Normal University Imaging Centre for Brain Research.

The samples from the UK Biobank consisted of 28,341 healthy individuals (13,727 males and 14,614 females). The exclusion criteria included any ICD-10 diagnosis of nervous system disease (field ID 41270), any ICD-9 diagnosis of nervous system disease (field ID 41271), algorithmically defined AD (field ID 42020), vascular dementia (field ID 42022), frontotemporal dementia (field ID 42024), motor neuron disease (field ID 42028), Parkinson’s disease (field ID 42032), progressive supranuclear palsy (field ID 42034), multiple system atrophy (field ID 42036), stroke (field ID 42006), ischaemic stroke (field ID 42008), intracerebral haemorrhage (field ID 42010), and subarachnoid haemorrhage (field ID 42012).

### Method 2: image acquisition and preprocessing

In the UK Biobank, images were acquired on 3T Siemens Skyra scanners using a magnetization-prepared rapid gradient echo (MPRAGE) sequence. The typical parameters included field of view (FOV) = 256 × 256 mm², matrix size = 256 × 256, slice thickness = 1 mm with no interslice gap, 208 slices, repetition time (TR) = 2000 ms, inversion time (TI) = 880 ms, and isotropic voxel size = 1 × 1 × 1 mm³. The detailed acquisition protocols are available at the UK Biobank documentation portal (https://biobank.ctsu.ox.ac.uk/crystal/crystal/docs/brain_mri.pdf).

For the ADNI dataset, T1-weighted images were acquired using vendor-specific 3D sequences harmonized across sites, including MPRAGE (Siemens), inversion recovery fast spoiled gradient recalled echo (IR-FSPGR; GE), and 3D turbo field echo (3D-TFE; Philips). In ADNI1&GO, scans were obtained at 1.5T with the following representative parameters: TR = 2400 ms, echo time (TE) = 3.5 ms, TI = 1000 ms, flip angle = 8°, FOV = 240 mm, matrix = 192 × 192, voxel size = 1.25 × 1.25 × 1.2 mm³, and 160 slices. In ADNI2 and ADNI3, most scans were acquired at 3T using accelerated protocols, with the following parameters: TR = 2300 ms, TE = 2.98 ms, TI = 900 ms, flip angle = 9°, FOV = 256 mm, matrix = 256 × 240, slice thickness = 1.2 mm, and voxel size = 1 × 1 × 1.2 mm³. Comprehensive protocol details are available at the ADNI documentation site (https://adni.loni.usc.edu/data-samples/adni-data/neuroimaging/mri/mri-scanner-protocols/).

For all T1-weighted MR images, standard preprocessing procedures in FreeSurfer^42^ were employed, which included nonuniformity correction, linear registration and skull stripping. The images were then resampled to a spatial size of 128×128×128 voxels with a resolution of 2 mm by using Advanced Normalization Tools (ANTs) (https://stnava.github.io/ANTs/).

### Method 3: network-specific brain age prediction model

The participants from the UK Biobank were randomly divided into the following three datasets: a training set (60%, n = 17,004), a validation set (20%, n = 5,668), and a test set (20%, n = 5,669) (Fig. 1a). On the basis of the Yeo-7 network atlas^24^, each T1-weighted MR scan was parcellated into 7 components, encompassing the VN, SMN, DAN, VAN, LIMN, FPN, and DMN. In this way, the features for each participant’s network were represented by the voxel intensities within the corresponding network regions.

Next, for each Yeo-7 network, a brain age model was trained via an improved SFCN model^25^ on the training set of the UK Biobank, a large-scale health reference cohort. The key advantage of the SFCN model lies in its ability to handle high-dimensional neuroimaging data. By leveraging fully convolutional layers, the SFCN model directly learns spatially distributed patterns of brain ageing without the need for manual feature extraction. Specifically, the architecture of the SFCN model consists of seven blocks as follows: the first five blocks each contain a 3×3×3 convolutional layer, batch normalization, ELU activation, and max pooling; the sixth block incorporates a 1×1×1 convolutional layer, batch normalization, ELU activation, and average pooling; and the final block includes a flattened layer, two fully connected layers with ELU activation, and a 50% dropout layer to prevent overfitting (Fig. 1b). This design enables the model to capture both global and localized brain features effectively for accurate age predictions.

### Method 4: estimation of individual network-based PADs

To assess the generalizability of the network-specific pretrained models, we applied them to unseen datasets from the UK Biobank and ADNI (Fig. 1c). Considering certain factors, such as regression dilution and non-Gaussian age distribution, which may cause biased predictions towards the average age of the cohort^43^, a linear correction was applied. For each sample with CA, an offset was calculated as follows:

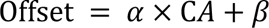

where *α* and *β* represent the slope and intercept, respectively, of a linear regression model of PAD (BA - CA) against CA in the training set^44^. This offset was subtracted from the raw BA estimate to obtain a corrected BA for each sample in the unseen data. Finally, the corrected PAD (corrected BA - CA) was subsequently computed and used for further statistical analysis.

### Method 5: diagnostic subgrouping for longitudinal analysis

In the nested case‒control design, we defined the following four diagnostic subgroups on the basis of baseline status, clinical progression, and data quality criteria: (1) normal control group (NC; n = 189), individuals without cognitive impairment at baseline and across all follow-ups; (2) sMCI group (*n* = 300), individuals who remained diagnosed with MCI throughout the follow-up period; (3) pMCI group (*n* = 157), individuals initially diagnosed with MCI who later progressed to AD; and (4) AD group (*n* = 276), individuals diagnosed with AD at baseline. Participants with diagnostic reversion (e.g., AD to MCI or MCI to NC) were excluded. To ensure adequate longitudinal coverage for tracking cognitive stability and maintaining statistical robustness, individuals in the NC and sMCI groups were required to have at least three assessments over a follow-up period of no more than six years, with a minimum interval of 12 months between visits.

### Method 6: longitudinal analyses of PADs in the AD continuum

We first fitted linear models to compare group-level differences in network-PADs at baseline. The models included group, age, sex, education level, and APOE status as fixed factors. Baseline PAD estimates were derived using the estimated marginal means (emmeans) package in R software (version 4.3.0)^45^. Overall group effects were evaluated using ANOVA, followed by pairwise comparisons of marginal means adjusted for multiple comparisons using Tukey’s HSD as a post hoc test.

Longitudinal trajectories of network-specific PADs were assessed with the use of a backwards timescale, where year 0 was defined as the year of AD diagnosis for converters and the end of follow-up for nonconverters. For each subgroup, a separate linear-effects model was fitted to estimate the trajectory of each network-specific PAD over time^46,47^. The models incorporated a linear function of retrospective time, with adjustments for group, covariates (e.g., age, sex, education level, and APOE status), and their interactions with time. Missing data were addressed via maximum likelihood estimation (MLE) during model fitting. Within-subject correlations were accounted for by correlated random intercepts and slopes of time. The annual rate of change for each individual was derived by combining the fixed-effect time slope, the group-specific time interaction, and the subject-specific random slope, thus accounting for heterogeneity in longitudinal trajectories.

To evaluate the significance of the group × time interaction, we performed an ANOVA to compare models with and without the interaction term, using Kenward-Roger’s degrees of freedom approximation^48^. Pairwise comparisons of marginal trends were conducted with Tukey-adjusted p values for multiple comparisons^45^. Finally, the fitted PADs for each network were integrated into a combined model to show the regional change sequence for the MCI to AD conversion.

### Method 7: biomarker and cognitive assessments

To characterize AD-related pathophysiology, we integrated genetic, fluid-based, and imaging biomarkers, along with standardized cognitive measures. The genetic variables included APOE ε4 carrier status and PHS, both of which were obtained from the ADNI database. Fluid biomarkers consisted of CSF Aβ_42_, tTau, and pTau181, which serve as molecular indicators of amyloid accumulation and tau-related neurodegeneration. The brain imaging markers included fluorodeoxyglucose positron emission tomography (FDG-PET) and flortaucipir (FTP) PET. Cerebral glucose metabolism was assessed using FDG-PET, with a global uptake index reflecting AD-related hypometabolism. Regional tau deposition was measured using FTP PET, with standardized uptake value ratios (SUVRs) extracted from the inferior temporal cortex, the entorhinal cortex, and a predefined meta-ROI. The tau meta-ROI was computed as the median uptake of voxels from the entorhinal, amygdala, parahippocampal, fusiform, inferior temporal, and middle temporal ROIs, normalized to the cerebellar crus.

Neuropsychiatric and cognitive status were assessed using the GDS, NPI, MMSE, and MoCA. All data were obtained from the Laboratory of Neuro Imaging (LONI) website (http://adni.loni.ucla.edu).

The PHS was calculated following the approach of Desikan et al. by integrating APOE status and 31 other genetic variants^49^. Detailed protocols for the measurement of CSF biomarkers have been described by Hansson et al.^50^, whereas those for PET tau imaging have been reported by Jack et al.^51^.

Additionally, procedures for deriving FDG-PET–based indices of cerebral glucose metabolism indices have been documented by Chen et al.^58^.

### Method 8: statistical analyses of PAD associations

A generalized linear model was utilized to separately examine the associations between baseline PADs and PAD change rates with the measurements within MCI. For the APOE genotype, age, sex, and education level were included as covariates, whereas for the other measures, the APOE genotype was additionally included as a covariate. All variables were normalized by the mean and standard deviation across all individuals before model fitting to ensure that the magnitude of the regression coefficient (*β*) was comparable across the models. Multiple comparisons were corrected via the Benjamini‒Hochberg method to control the false discovery rate (FDR), with a corrected α level of 0.05.

### Method 9: mediation analyses of biomarkers, brain age, and cognition

Given the dynamic interplay among cognitive function, genetic factors, pathological characteristics, and brain systems, we hypothesized that AD risk genes and pathology influence cognitive performance by selectively modulating the ageing of specific brain networks. To test whether baseline PADs and their annual change rates mediate the relationship between AD-related biomarkers and cognitive performance (both baseline scores and longitudinal changes), we performed mediation analyses using two-step ordinary least squares (OLS) regression models^54^. For each biomarker–PAD–cognition triplet, we first modelled the effect of the biomarker on the PAD (mediator). In the second step, we assessed the joint effects of both the biomarker and PAD on cognitive performance. All models were adjusted for age, sex, and years of education as covariates and were fitted to a subset of data with complete observations.

Indirect effects were estimated as the product of the two regression coefficients and evaluated using a nonparametric bootstrap procedure with 5,000 resamples to derive 95% confidence intervals. Statistical significance was determined by intervals excluding zero. To correct for multiple comparisons across mediation pathways, the Benjamini–Hochberg method was used to control the FDR rate at 5%.

### Method 10: machine learning model for predicting the MCI to AD conversion

We implemented a random forest classifier to predict whether individuals with MCI progress to AD. Model evaluation was conducted using 10-fold cross-validation, which was repeated 10 times, to ensure robust performance estimation. To assess the incremental predictive value of network-based brain ageing features, we conducted a feature ablation analysis by progressively enriching the input feature set across the following three models:

- Model 1 (benchmark), genetic and pathological biomarkers only;
- Model 2, Model 1 plus baseline PADs;
- Model 3, Model 2 plus short-term longitudinal PAD change rates.

Individual PAD change rates were derived from longitudinal data from baseline and at a second time point within six years prior to AD diagnosis. Model performance was assessed using accuracy, recall, specificity, F1 score, and AUC-ROC, with the results reported as the means ± standard deviations across repetitions. A fixed random seed ensured reproducibility. To test the statistical significance of the model improvements, we conducted two-sided paired t tests on the AUC‒ROC values between successive models.

## Data availability

Access to UK Biobank data is available upon request through its official website (https://www.ukbiobank.ac.uk/), which is subject to its data access policies and approval procedures. Similarly, ADNI data can be accessed by qualified investigators upon request, pending approval from the ADNI Data and Publications Committee. Detailed instructions for submitting a request are provided at https://adni.loni.usc.edu/data-samples/access-data/.

## Code availability

Analyses were performed using R software (v4.3.0) and Python (v3.10.11). Model training and prediction code is available at https://github.com/xuanmer/NetworkBrainAge, and code for subsequent statistical analyses is available at https://github.com/xuanmer/StatisticalAnalysis.

## Notes

### Competing Interest Statement

The authors have declared no competing interest.

### Author Declarations

The Ethics Committee of the Institutional Review Board at the Beijing Normal University Imaging Centre for Brain Research.

